# Hypertension increases PPV for polycystic kidney disease in *PKD1* and *PKD2* variant carriers

**DOI:** 10.1101/2023.10.30.23297693

**Authors:** Natalie Telis, Lisa McEwen, Alexandre Bolze, Daniel P. Judge, Pamala A. Pawloski, Joseph J. Grzymski, Catherine Hajek, Kelly M. Schiabor Barrett, Nicole L. Washington, Elizabeth T. Cirulli

## Abstract

Autosomal dominant polycystic kidney disease (ADPKD) is the leading genetic form of kidney disease. Although rare causal variants in the *PKD1* and *PKD2* genes have been identified, their penetrance and the disease progression and outcome are known to vary widely in carriers, and treatment efficacy in these carriers lags compared to patients with chronic kidney disease (CKD). We characterize the presentation and progression of ADPKD by identifying variant carriers in a novel exome sequencing cohort as well as in the UK Biobank. We show that the positive predictive value of hypertension for future diagnosis of kidney disease in variant carriers is extremely high: 74% and 66% for *PKD1* and *PKD2* respectively. It is also highly preemptive, with hypertension occurring an average of 11 years before a kidney disease diagnosis. We estimate measures of kidney function for individuals at timepoints prior to their diagnoses and find that *PKD1* and *PKD2* variant carriers show significantly decreased eGFR an average of 5 years before their eventual diagnosis of kidney disease. In fact, at a standard assessment performed a mean of 5 years prior to kidney disease diagnosis, 54% of *PKD1* and *PKD2* variant carriers with hypertension already meet the diagnostic threshold for kidney disease, and their eGFR levels are statistically indistinguishable from carriers who have already been diagnosed. Together, these findings suggest that ADPKD could be anticipated and treated significantly earlier using a combination of targeted sequencing and routine monitoring.

## Introduction

Kidney disease is common in the developed world and is a major cause of morbidity and mortality in the United States. Autosomal dominant polycystic kidney disease (ADPKD) is the most common genetic form of kidney disease, affecting approximately 1 in 500 to 1 in 1,000 individuals worldwide (1). The majority of genetic ADPKD cases are due to mutations in the gene *PKD1*, with a smaller percentage attributed to *PKD2*. These genes encode polycystins which form a complex involved in the formation and development of renal tubules. The clinical manifestation of ADPKD arises from the development and expansion of cysts within the renal parenchyma, leading to a constellation of symptoms that can impact quality of life and overall health. Most patients present with at least two kidney cysts by age 40 (1). In contrast, chronic kidney disease (CKD) does not typically present with cysts. However, in both diseases, damage to the kidneys results in hypertension, which leads to cardiovascular damage and morbidity, which further damages the kidneys and other tissues (2).

While kidney damage is generally irreversible, cardiovascular and other comorbidities can be treated, and the rate of kidney damage can be modulated. The progression of disease can be slowed by decreasing the risk of acute kidney damage due to UTIs or other infections as well as by managing hypertension and other forms of cardiovascular morbidity (3); (4). However, as kidney disease progresses and organ function declines, patients eventually reach end-stage renal disease (ESRD) and typically require renal replacement therapy (RRT). The age at which RRT is initiated and the 5 year life expectancy has increased steadily over the last 30 years for individuals with CKD, indicating improvement in lifespan due to the efficacy of new treatments (5–7). However, the age for RRT in patients with ADPKD has not changed over the same time period.

There may be many reasons that treatment for ADPKD has not improved. Although causal mutations in *PKD1* and *PKD2* are well characterized, the disease presentation and penetrance of *PKD1* and *PKD2* mutations is highly variable. Onset and progression of ADPKD varies significantly by which gene is mutated as well as amongst individuals with mutations in the same gene, and is even variable for affected individuals within the same family ((8–10). Additionally, the timing of onset and diagnosis of ADPKD is somewhat complicated by the significant delay between the onset of renal cyst formation and the manifestation of symptoms of kidney disease. In ADPKD, development of cysts in the kidneys can start very early, including in utero, but even a serious increase in kidney volume in childhood may not cause a decline in kidney function (11,12). While hypertension occurs prior to disease in a majority of ADPKD patients and is associated with a significantly worse prognosis, it can sometimes be found in *PKD1* and *PKD2* carriers in childhood without signs of kidney damage (13).

Since existing treatments can only reduce the rate of kidney decline and not reverse extant damage, it is critical to understand the point at which kidney damage has begun to ensure prompt and effective disease management, mitigate the harms of kidney damage, and prolong life. To understand the progression of ADPKD and inform a potential genetic screening strategy, we identify individuals with putative loss of function mutations in *PKD1* and *PKD2* in the UK Biobank (UKB) and in individuals sequenced at Helix. We then characterize their clinical presentation of kidney disease by looking at enrichment and time of onset for diagnosis of hypertension, kidney disease, and end-stage renal disease. In the UKB, a uniform time of assessment at which blood labs were collected enables us to assess *PKD1* and *PKD2* LoF carriers’ kidney function and other subclinical characteristics of disease presentation at various stages in the disease progression. This enables us to develop a quantitative characterization of the timing and progression of ADPKD, and understand the material consequences on kidney function directly at various stages of disease, including prior to diagnosis.

## Methods

### Subjects and genetic data

We utilized the UKB population level exome OQFE pVCFs for 469,811 individuals (field 23157, with genotypes set to missing when DP<7 for SNVs and <10 for indels, and variants excluded if there were no homozygotes or the max allelic balance was <0.15 for SNVs or <0.2 for indels as per (14) as well as the imputed genotypes from GWAS genotyping (field 22801-22823). The UKB study was approved by the North West Multicenter Research Ethics Committee, UK. For UKB, we were able to divide the cohort into prospective and retrospective diagnoses by dividing the time at first incidence of phenotype using the date of the baseline assessment visit.

We also utilized 62,406 samples that were sequenced and analyzed at Helix using the Exome+^®^ assay as previously described, recruited from the Healthy Nevada Project (n=37,989, sequenced Jan 2018 to Mar 2023); myGenetics (n=15,104, sequenced May 2022 to Mar 2023); and In Our DNA SC (n=9,313, sequenced Dec 2021 to Mar 2023) (Supplementary Table 1) (15). The Helix cohort studies were reviewed and approved by their applicable Institutional Review Boards (IRB, projects 956068-12 and 21143). All participants gave their informed, written consent prior to participation. All data used for research were deidentified.

### Phenotypes

UKB data were provided from the UKB resource (accessed September 2022). ICD codes and associated dates (both ICD-9 and ICD-10) were collected from inpatient data (category 2000), cancer register (category 100092) and the first occurrences (category 1712), which records the earliest instance of a diagnosis from the Primary Care data, Hospital inpatient data, Death Register records, and self-reported medical conditions mapped to a ICD10 code at a three character resolution (i.e., E11 instead of E11.0). Helix cohort phenotypes were processed from Epic/Clarity Electronic Health Records (EHR) data as previously described and updated as of January 2023 (15). International Classification of Diseases, Ninth and Tenth Revision ICD codes and associated dates (ICD-9 and ICD-10-CM) were collected from available diagnosis tables (from problem lists, medical histories, admissions data, surgical case data, account data, claims, and invoices). All datasets were transformed into the OMOP Common Data Model v5.4. Therefore, diagnosis codes were mapped to SNOMED terms and represented by concept ids. To be comprehensive in our cohort definition, we used both the source concept id and the standard concept id to extract relevant diagnoses.

Hypertension, kidney disease and end-stage renal disease (ESRD) were defined by phecode review as well as a literature search around clinical diagnosis codes (ICD10, ICD10CM, ICD9, SNOMED) given the different natures of the source datasets. Because all diagnoses were mapped to SNOMED, we identified the first record of each included SNOMED term to represent the first occurrence of disease for the associated individual. A complete list of diagnosis codes and mapped SNOMED terms corresponding to each phenotype (hypertension, kidney disease and ESRD) can be found in Supplementary Tables 2, 3 and 4, respectively.

Blood biomarkers and lab values were sourced from the UKB fields 30500-30900. We calculate eGFR based on the CKD-EPI equation for all individuals with a measured serum creatinine (field 30700) (16).

The UKB recruitment assessment is a timepoint affected only by participation bias in the UKB rather than the independent experience of a symptom and as a result gives us the opportunity to evaluate biomarkers measured at this time point, rather than labs ordered to perform a differential diagnosis based on observed symptoms. The median age at assessment is 58.5 years (mean = 57.2 + 8.1 years) with a median of 8.5 years of followup (mean = 7.25 + 4.5 years), allowing us to analyze nearly a decade of prospective diagnosis data after an initial biomarker measurement. To classify prospective and retrospective cases, we compared the assessment date and the first diagnosis date. The first diagnosis date is the date on the earliest incidence of any of the SNOMED terms included in the UKB cohort in our list of phenotypes. The assessment date is the date when the biobank sample and survey were taken (field 53). Given this assessment date, we defined people as prospective or retrospective cases for each phenotype we measured.

#### Annotation and qualifying *PKD1* and *PKD2* variants

Variant annotation was performed with VEP-104(17). Coding regions were defined according to Gencode version GENCODE 33 using the MANE / Ensembl canonical transcript ENST00000262304.9 (*PKD1*) and ENST00000237596.7 (*PKD2*) to determine variant consequence (18,19). Variants from the exome were restricted to CDS (coding sequence) regions plus essential splice sites. Genotype processing for Helix data was performed in Hail 0.2.115-10932c754edb (20). Variants were classified as LoF if the consequence was stop_lost, start_lost, splice_donor_variant, frameshift_variant, splice_acceptor_variant, or stop_gained. Additionally, LoF variants were filtered to MAF < 0.1% in each gnomAD and local UKB or Helix population and, based on our prior study of these genes and phenotype with the Power Window method (21), fewer than 20 carriers in UKB. Any individual with a variant meeting this criteria was determined to be a carrier of either a *PKD1* or *PKD2* LoF variant, and no individuals in our study had a qualifying variant in both genes.

#### Statistical analyses

Statistical analyses were run using the statsmodel package in Python 3.7.3. For binary variables, logistic regression or Firth logistic regression was used; for quantitative variables, linear regression was used after rank-based inverse normal transformation. For time to event analyses, the lifelines package was used, and the most recent current age was considered to be the highest of either the age at the most recent diagnosis the participant received anywhere in their medical record, or their assessment age (22). The main analyses were performed on all ancestries together because we are analyzing very rare variants (MAF<0.1%) collapsed together, as previously described, and the associations described perform similarly when restricted to a European-only cohort with principal components included as covariates (15,23,24).

## Results

### *PKD1* and *PKD2* carriers significantly enriched for diagnoses of kidney disease

To understand the prevalence of *PKD1* and *PKD2* LoF variants in the general population, we analyzed 469,811 exome sequences from the UKB and 65,934 from Helix cohorts. We identified 124 individuals with rare *PKD1* LoF variants and 94 with rare *PKD2* LoF variants in the UKB (a combined prevalence of 0.049%). We identified a similar prevalence (0.047%) in the Helix cohorts, with 31 *PKD1* LoF variant carriers and 11 *PKD2* LoF variant carriers.

In the UKB, we find the incidence of kidney disease (KD) to be 47% in *PKD1* carriers and 48% in *PKD2* carriers, compared to 6% in the population of non-carriers. In the Helix cohorts, these values were 42% for *PKD1* carriers, 36% for *PKD2*, and 7% for non-carriers. The risk of kidney disease increases with age, and the average age of the Helix cohort is quite a bit younger than the average age of the UKB cohort (53.6 vs 64.5; t-test, p < 2.2e-16), so to ensure we capture effects on prevalence driven by age, we also compare prevalence in younger and older individuals (see Methods). In both cohorts, the age at disease onset varies significantly by gene; 84% of *PKD1* LoF carriers who will develop kidney disease are diagnosed with kidney disease by age 60, as compared to 51% of *PKD2* LoF carriers (Kolmogorov-Smirnov, p = 5.3e-4) and 18% of non-carriers (Kolmogorov-Smirnov, p=< 2.2e-16).

### Hypertension incidence has significant positive predictive value for onset of kidney disease

A typical step in the diagnostic pathway for identifying kidney disease is a finding of hypertension, which is enriched in individuals with kidney disease (25). The incidence of hypertension in non-carriers in the UKB cohort is 40%, and only 35% of these cases have onset before the age of 60 (see Supplementary Fig. 1). In comparison, the incidence of hypertension in *PKD1* and *PKD2* carriers is elevated at younger ages. In the UKB, 62% of *PKD1* variant carriers have hypertension, significantly more than non-carriers (binomial test, p = 0.0002), and 91% of carriers diagnosed are diagnosed before age 60. Similarly to *PKD1* carriers, 57% of all *PKD2* carriers have hypertension and 78% of carriers diagnosed are diagnosed before the age of 60, which is significantly greater than the rate in non-carriers (binomial test, p = 0.009).

While hypertension is often the beginning of the discovery and diagnosis of kidney disease, the incidence of kidney disease in non-carriers with hypertension is only 12%, making hypertension an unreliable predictor of eventual kidney disease in non-carriers, with low PPV. However, the PPV for kidney disease is much higher in LoF carriers with hypertension: 75% for *PKD1* and 64% for *PKD2*. Because of this high PPV and a relatively lower incidence of kidney disease in carriers without hypertension–2% for *PKD1* and 33% for *PKD2*–the diagnosis of hypertension is a much more clinically meaningful warning sign in these individuals. We find similar results in the Helix cohorts, where 75% of *PKD1* carriers and 50% of *PKD2* carriers with hypertension have kidney disease, compared to 1 out of the 3 *PKD1* carriers without hypertension and 0 out of the 3 *PKD2* carriers without hypertension (see Supplemental Table 5). This difference is not explained by the increasing incidence of kidney disease at older ages (see Supplemental Table 6).

### Timing of kidney disease onset and progression

To more deeply characterize these differences in disease onset, presentation and progression, we examine the timeline of disease progression by comparing the onset of hypertension, kidney disease, and end-stage renal disease (ESRD). We identify the timeline of onset by inspecting the inpatient data (see Methods) to get a precise age for each individual at the onset of hypertension, kidney disease, and/or end-stage renal disease. The onset of hypertension in *PKD1* variant carriers far precedes the onset in both *PKD2* carriers and non-carriers (Fig. 2a) (Kolmogorov-Smirnov, p = 0.0001 and < 2.2e-16 respectively). 30% of *PKD1* carriers have hypertension by age 40, compared with 11% of *PKD2* carriers (binomial test, p = 2.7e-9) and only 3.1% of non-carriers (binomial test, p < 2.2e-16). Comparatively, it takes until age 50 for 30% of *PKD2* carriers to develop hypertension and until age 64 for 30% of non-carriers to develop hypertension. Notably treatment for hypertension is not associated with either age of onset of hypertension or rapidity of progression of disease (see Supplementary Fig. 2).

**Figure 1.**
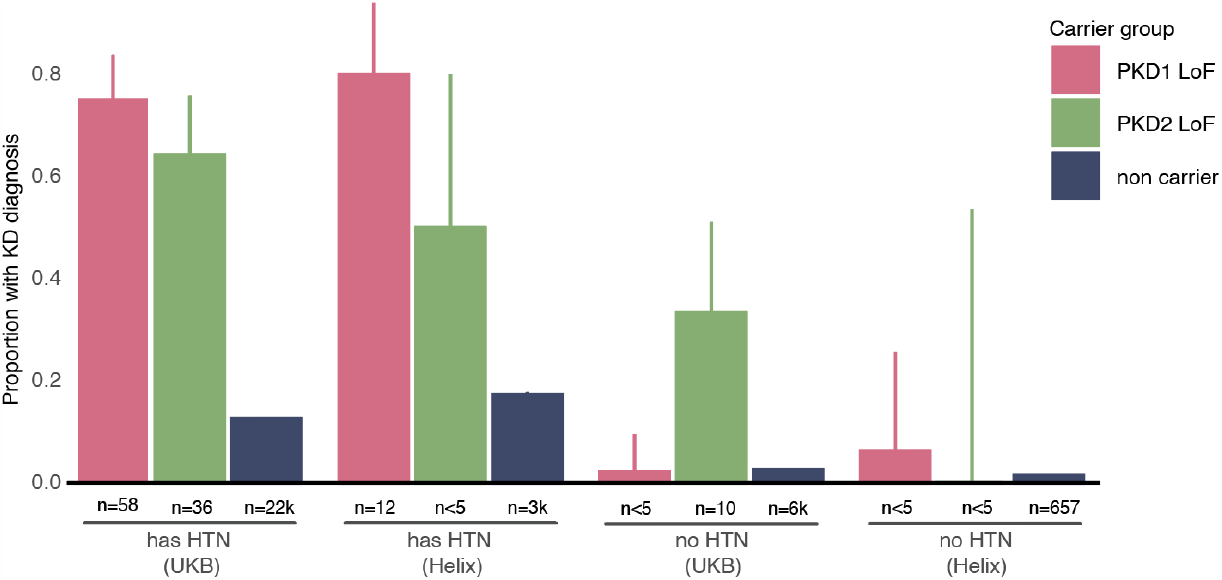
Positive predictive value of hypertension on diagnoses of kidney disease in *PKD1* and *PKD2* LoF carriers. Proportion of kidney diagnosis for individuals with hypertension (UKB), with hypertension (Helix cohorts), without hypertension (UKB) and without hypertension (Helix cohorts). Individuals are classified as either having a *PKD1* LoF variant, a *PKD2* LoF variant, or no known variants. Number of individuals diagnosed in each category is displayed beneath the bar. Error bars for proportions are calculated by calculating the binomial errors and are present for all groups (although miniscule for the large groups of non-carriers).

**Figure 2.**
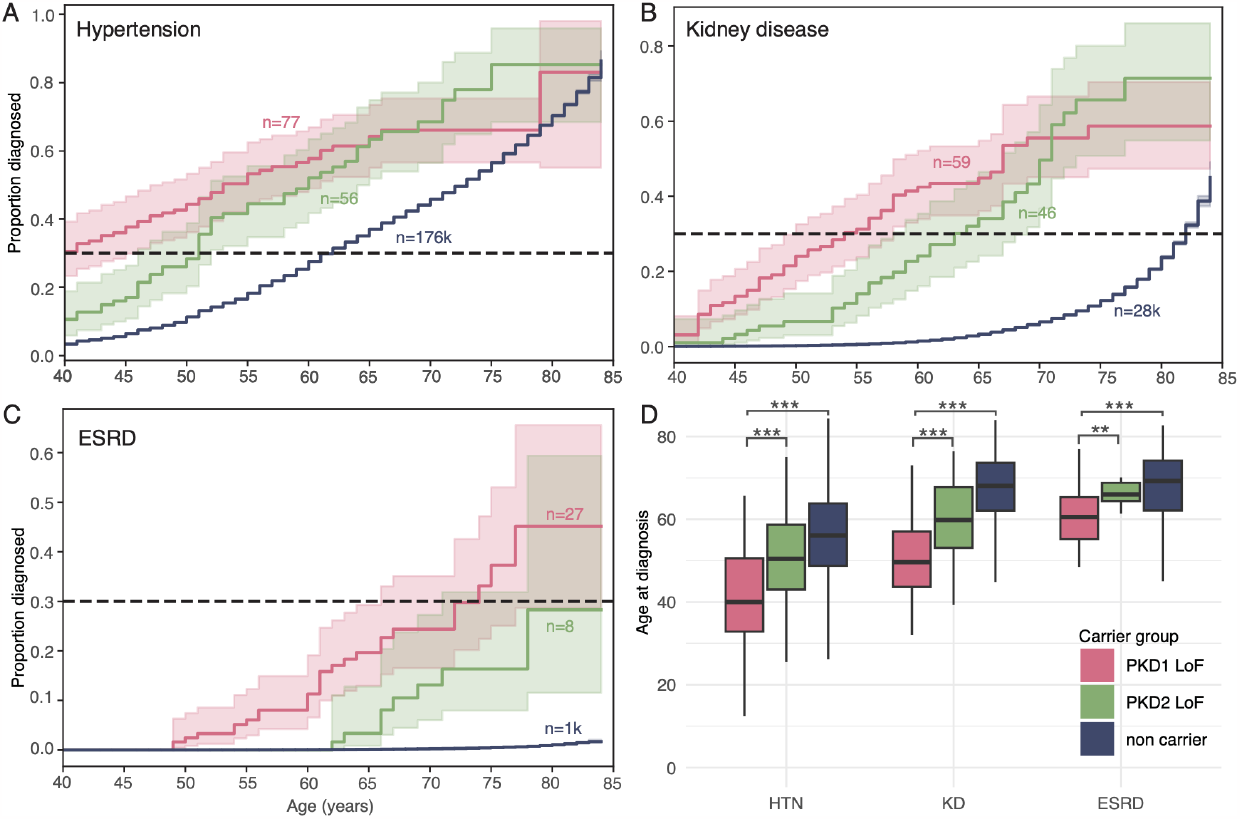
Hypertension and kidney disease age at onset is significantly earlier for *PKD1* and *PKD2* LoF carriers. Proportion of individuals with a diagnosis of **(A)** hypertension, **(B)** kidney disease, or **(C)** end-stage renal disease by a given age. Number of individuals included in each line is labeled. A proportion of 0.3 (30%) is marked on the graph as a point of comparison and consistency. **(D)** Age at first diagnosis of hypertension (HTN, left), kidney disease (KD, middle), or end-stage renal disease (ESRD, right). Colors represent whether individuals carry a *PKD1* or *PKD2* LoF variant or if they are noncarriers. Asterisks denote levels of significance as determined by a t-test. * p = <0.05, ** p = <0.01, *** p = < 0.001.

After onset of hypertension, the progression to kidney disease is rapid for *PKD1* carriers. We examine the timing of onset of kidney disease relative to this baseline age at which 30% of individuals have been diagnosed with hypertension. Of the *PKD1* carriers diagnosed with hypertension by age 40, 50% have been diagnosed with kidney disease by age 48 (see Supplementary Fig. 3). The onset and progression is substantially slower in *PKD2* LoF carriers: In addition to the difference in baseline age at hypertension diagnosis (50% by age 40 for *PKD1* vs. 50% by age 50 for *PKD2*), it also takes 18 more years (until age 68) for 50% of PKD2 carriers to be diagnosed with kidney disease (Fig. 2b). These ages differ significantly from the progression of noncarriers from their comparatively much older baseline; only 9.6% of those diagnosed with hypertension by age 64 (the 50% diagnosis age) developed any form of kidney disease in the 10 years that followed (up to 12% with 20 years follow up). However, the mean number of years between a hypertension and kidney disease diagnosis is fairly similar in all groups; 11 years for non-carriers and 12 years for *PKD1* and *PKD2* variant carriers (t-test, p = 0.35).

End stage renal disease is fairly rare in *PKD1* and *PKD2* variant carriers in the UKB dataset, perhaps because of known participation bias affecting the representation of particularly adverse health outcomes (26). In spite of this, the mean age at ESRD in UKB is 60 years old for *PKD1* carriers and 67 years old for *PKD2* carriers, which is similar to estimates from the literature (see e.g. (27,28). Variant carriers who will develop ESRD are diagnosed with hypertension slightly earlier than those who will not (mean age = 41.4 versus 46.4 years old, Kolmogorov-Smirnov test, p = 0.04, despite similar followup time scales). In *PKD1* and *PKD2* LoF carriers diagnosed with hypertension before the age of 45, 36% will develop ESRD, compared with 17% of those diagnosed with hypertension after the age of 45 (binomial test, p = 0.0009).

### Prospective identification of future disease

At assessment (median age 58.5 years), all UKB participants are measured for a standard set of blood labs, providing a snapshot into the health of each person in a manner not captured in standard medical care. Based on the date of assessment, we classify individuals as having either prospective diagnosis of disease (after assessment, during the median of 8.5 years of followup) or retrospective diagnosis (before assessment; for more, see Methods). Of approximately 180k hypertension diagnoses, 69% occurred before the assessment date; however, only 23% of all kidney disease diagnoses occur before the assessment date, allowing us to examine a large cohort of individuals in an ostensibly pre-kidney-disease state.

An eGFR from 60-90 is an early warning sign of kidney disease, with an eGFR of less than 60 indicating a diagnosis of kidney disease. At assessment, eGFR is significantly lower in carriers with kidney disease compared to carriers without kidney disease, regardless of whether they are a prospective or retrospective case (t-test, p < 2.2e-16; see Fig. 3). Strikingly, eGFR is also significantly lower in all carriers with kidney disease compared to non-carriers with kidney disease (t-test, p = 1.7e-11), especially in prospective cases (p=7.8e-9) and weakly in retrospective cases (p = 0.01). This is not true for *PKD1* and *PKD2* carriers who never get a kidney disease diagnosis in the observation period, regardless of whether they have hypertension (see Supplementary Fig. 4).

**Figure 3.**
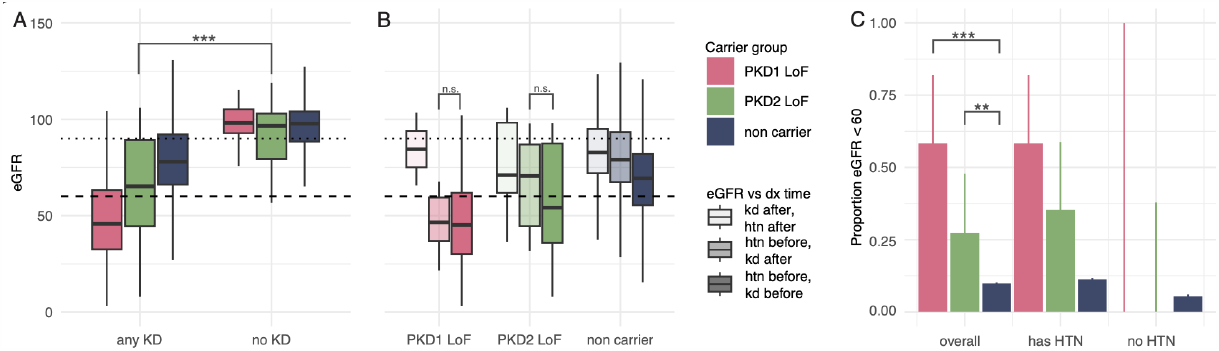
In UKB, *PKD1* and *PKD2* LoF carriers with a prospective diagnosis of kidney disease show signs of disease at least 5 years before diagnosis. **(A)** eGFR calculated with the CKD-EPI equation (16) for (left) individuals with any KD diagnosis recorded in the UKB medical record dataset and (right) individuals with no KD at any point before the assessment blood draw or in the observation period since then. **(B)** eGFR calculated as before for only individuals with a hypertension and KD diagnosis, divided by carrier group and timing of the assessment blood labs relative to the diagnosis of hypertension and kidney disease. From left to right: *PKD1* LoF carriers, *PKD2* LoF carriers, and non carriers with HTN and KD, divided into diagnoses timing: hypertension before and kidney disease after assessment; hypertension before and kidney disease before assessment; hypertension and kidney disease both after assessment. Lower thick dashed line represents the diagnostic threshold of KD (eGFR < 60). Upper light dashed line represents early stage KD (eGFR < 90). **(C)** Proportion of individuals with eGFR below a clinical diagnostic threshold (<60) who do not have a diagnosis of kidney disease at assessment, summarized overall, and divided by the presence or absence of a hypertension diagnosis. Asterisks denote levels of significance as determined by a Kolmogorov-Smirnov test. * p = <0.05, ** p = <0.01, *** p = < 0.001.

This statistically significant difference in eGFR levels has clinical implications. At the assessment timepoint, 38% of all *PKD1* or *PKD2* variant carriers who will be diagnosed in the future already have an eGFR below the diagnostic threshold (60), compared with just 9.8% of non-carriers who will be diagnosed (binomial test, p = 3.8e-6). This difference is strongest in *PKD1* variant carriers; 58% of *PKD1* variant carriers who go on to get a diagnosis already have an eGFR below the diagnostic threshold at assessment (binomial test, p= 2.5e-5). Additionally, 27% of *PKD2* variant carriers who go on to get a diagnosis meet the diagnostic threshold at assessment, which is also significantly more than non-carriers (binomial test, p = 0.01).

Hypertension also has a high PPV for the incidence of an early stage low eGFR. While 54% of all *PKD1* LoF carriers have an eGFR < 90, this increases to 72% of *PKD1* LoF variant carriers with hypertension but is only 23% in *PKD1* LoF carriers without hypertension. Likewise, while 44% of individuals with a *PKD2* LoF variant have an eGFR < 90, this increases to 61% in individuals with hypertension and is 26% in individuals without hypertension.

Although carriers with prospective cases of kidney disease are diagnosed an average of 5 years after the initial timepoint, their lab values do not differ significantly from those diagnosed before the initial timepoint, suggesting that many individuals are being diagnosed later than they should be. Notably, of individuals who are undiagnosed at the assessment date, 73% meet a diagnostic criteria for early kidney disease (eGFR < 90) and 38% meet the diagnostic criteria for kidney disease (eGFR < 60). In short, a majority of *PKD1* and *PKD2* LoF carriers have notable kidney damage well in advance of their diagnosis with kidney disease and a plurality already meet existing guidelines for diagnosis and treatment.

## Discussion

*PKD1* and *PKD2* variants are the major known genetic contributors to autosomal dominant polycystic kidney disease. In our study we examine the differences in disease presentation at a clinical and biomarker level of patients with a rare LoF variant in *PKD1* or *PKD2*. We show that having a rare LoF variant is strongly associated with the onset of the stages of kidney disease. Consistent with the literature, we observe that *PKD1* carriers have a significantly earlier onset of disease than *PKD2* carriers, beginning with the much earlier incidence of hypertension and transitioning in much greater numbers to kidney disease and to end-stage renal disease (ESRD). Importantly, we find that hypertension clearly provides an early warning sign of kidney disease progression in *PKD1* and *PKD2* carriers. In both, it precedes diagnosis of kidney disease by an average of at least 11 years with a PPV of 71%. Additionally, earlier diagnoses of hypertension are associated with increased incidence of severe morbidity.

While general risk factors for developing kidney disease and for progression are well understood, the timing of disease start and point of effective intervention in ADPKD has remained unclear. Because many kidney disease cases in the UKB are diagnosed after the assessment visit and measurement of blood lab values, our study was able to divide kidney disease cases into prospective and retrospective case cohorts to understand the relationship between risk factors, early symptoms, and the onset of kidney dysfunction.

For individuals with no future diagnosis of kidney disease, kidney function in carriers with hypertension does not appear altered from non-carriers with hypertension. However, carriers with prospective cases of kidney disease have decreased eGFR at least five years before their kidney disease diagnosis. While this is true for non-carriers as well, *PKD1* and *PKD2* variant carriers are disproportionately underdiagnosed at the assessment time. Based on their eGFR, 83% of *PKD1* LoF carriers with hypertension would meet diagnostic criteria for at least early stage kidney disease at assessment and 53% would meet diagnostic criteria for kidney disease disease. Most importantly, undiagnosed carriers are not distinguishable on metrics of kidney function from carriers with a diagnosis prior to assessment (*PKD1* carriers = 47.8 (p = 0.71); *PKD2* carriers = 68.2 (p = 0.08); see Fig. 3A).

Kidney disease treatment is about decelerating damage and comorbidity reduction, and it is clear from our prospective study that kidney function decline in *PKD1* and *PKD2* variant carriers has begun well before their diagnosis with kidney disease. This is also true in non-carriers. However, less than 10% of non-carriers meet diagnostic criteria at the assessment, compared to a majority of *PKD1* and *PKD2* carriers. While monitoring eGFR in the population at large would also identify some individuals with decreased kidney function, it has a significantly greater positive predictive value in individuals who carry a *PKD1* or *PKD2* variant. Monitoring eGFR in *PKD1* and *PKD2* carriers, especially those who have already developed hypertension, would be a highly specific way to screen for the onset of kidney disease. If all individuals who develop hypertension prior to age 40 were assessed for LoF variants in *PKD1* and *PKD2*, then 1/320 would be carriers (see Supplementary Fig. 4), a prevalence similar to that seen for other genetic conditions like Lynch syndrome, and not far from the American College of Medical Genetics & Genomics (ACMG) guidelines for genetic screening (29,30).

Notably, there are a number of individuals who appear resilient to disease. Although the mean age at ESRD for *PKD1* variant carriers is estimated to be between 54-58 (27,28), 34% of individuals carrying *PKD1* variants in our cohort are entirely unaffected by kidney disease even after age 60. Interestingly, only 37% of these unaffected carriers have hypertension, compared to 50% of non-carriers over the age of 60. In our analysis we did not find LoF variants carried by these individuals to be distinguishable in these resilient individuals in terms of the type of LoF variant or gene position, suggesting that further studies may identify other genetic or environmental factors affecting penetrance in these individuals.

## Data Availability

Summary data produced in the present work are contained in the manuscript.

## Acknowledgments

This research has been conducted using the UK Biobank resource under application number 40436. This work uses data provided by patients and collected by the NHS as part of their care and support, re-used with the permission of the UK Biobank under Application Number 40436. Copyright © (2023), NHS England. All rights reserved. This research also used data assets made available by National Safe Haven as part of the Data and Connectivity National Core Study, led by Health Data Research UK in partnership with the Office for National Statistics and funded by UK Research and Innovation (grant ref MC_PC_20058). Funding was provided to DRI by the Nevada Governor’s Office of Economic Development. Funding was provided to the Renown Institute for Health Innovation by Renown Health and the Renown Health Foundation. We acknowledge the entire Helix Bioinformatics team for their contributions to the production exome sequencing pipeline. We thank all of the genomic representatives of the Healthy Nevada Project, myGenetics,and In Our DNA SC (Helix Cohorts). We thank Renown Health, DRI, Medical University of South Carolina, and HealthPartners for helping to launch the Helix Cohorts.

## Ethics Declaration

The Helix cohorts were reviewed by Salus IRB (Reliance on Salus for all sites) and approved (approval number 21143), the WIRB CG IRB (Western Institutional Review Board, WIRB-Copernicus Group) and approved (approval number 20224919), the MUSC Institutional Review Board for Human Research and approved (approval number Pro00129083), and the University of Nevada, Reno Institutional Review Board and approved (approval number 7701703417). The UK Biobank study was approved by the North West Multicenter Research Ethics Committee, United Kingdom. All participants gave their informed, written consent before participation. All data used for research were de-identified.

## Tables

**Table 1.** Summary of UK Biobank and Helix cohorts incidence of *PKD1* and *PKD2* variants.

|  | UKB |  | Helix |  |
| --- | --- | --- | --- | --- |
|  | Non-carrier | *PKD1* & *PKD2* LoF | Non-carrier | *PKD1* & *PKD2* LoF |
| N (%) | 440,945 (99.9%) | 210 (0.04%) | 65,904 (99.9%) | 31 (0.04%) |
| Mean current age (SD) | 64.5 (9.9) | 63.9 (9.5) | 53.7 (16.7) | 48.6 (15) |
| N female (%) | 199,969 (54%) | 91 (58%) | 45,672 (68%) | 25 (80%) |

**Table 2.** Hypertension ICD10 codes and matching SNOMED code.

| ICD10 code | Matching SNOMED |
| --- | --- |
| I10 | 59621000 |
| I11 | 64715009 |
| I11.0 | 5148006,46113002 |
| I11.9 | 60899001 |
| I12 | 38481006,104931000119100 |
| I12.9 | 38481006,104931000119100 |
| I12.0 | 49220004 |
| I13 | 86234004,8501000119104 |
| I13.0 | 194779001 |
| I13.1 | 194780003,8501000119104 |
| I13.2 | 194781004,8501000119104 |
| I13.9 | 86234004 |
| I15 | 31992008 |
| I15.0 | 123799005 |
| I15.1 | 28119000 |
| I15.2 | 194788005 |
| I15.8 | 31992008 |
| I15.9 | 31992008 |

**Table 3.** Kidney disease ICD10 codes and matching SNOMED codes.

| ICD10 code | Matching SNOMED |
| --- | --- |
| N18 | 709044004 |
| N18.1 | 431855005 |
| N18.2 | 431856006 |
| N18.3 | 433144002 |
| N18.4 | 431857002 |
| N18.5 | 433146000 |
| N18.6 | 46177005 |
| N18.8 | 90688005 |
| N18.9 | 709044004 |
| Q61 | 722223000 |
| Q61.0 | 369071000119105,5941000119101 |
| Q61.1 | 28770003 |
| Q61.2 | 765330003 |
| Q61.3 | 82525005 |
| I12 | 38481006,104931000119100 |
| I12.9 | 38481006,104931000119100 |
| I12.0 | 49220004 |
| I13 | 86234004,8501000119104 |
| I13.0 | 194779001 |
| I13.1 | 194780003,8501000119104 |
| I13.2 | 194781004,8501000119104 |
| I13.9 | 86234004 |
| I15 | 31992008 |
| I15.0 | 123799005 |

**Table 4.** End-stage renal disease (ESRD) ICD10 codes and matching SNOMEDs.

| ICD10 code | Matching SNOMED |
| --- | --- |
| N18.5 | 433146000 |
| N18.6 | 46177005 |

**Table 5.**
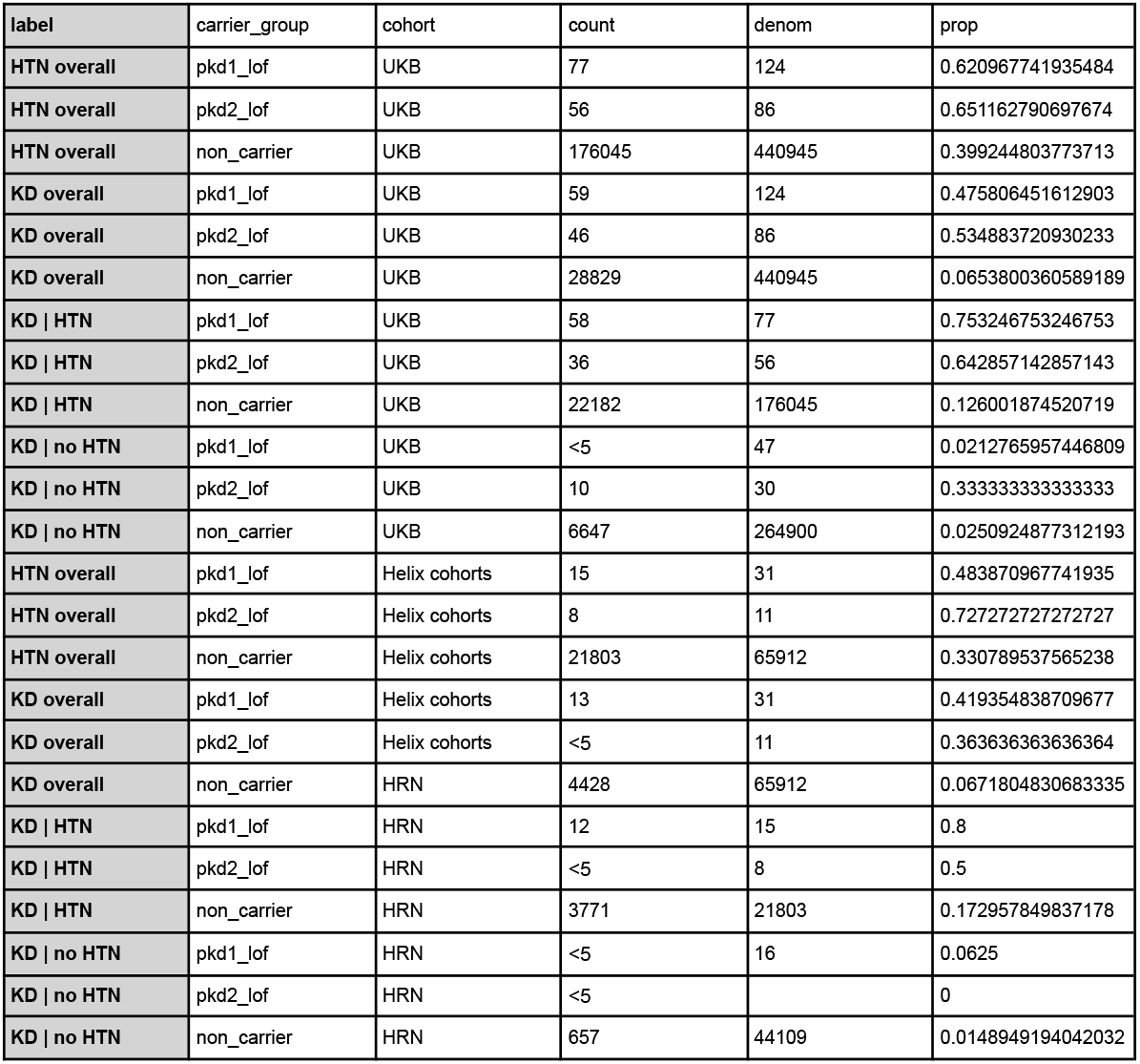
Counts of individuals classified into each diagnosis label (‘label’) divided by carrier group (PKD1, PKD2 variant, or non carrier) for both UKB and Helix cohorts.

**Table 6.** % individuals diagnosed with hypertension by decade of age.

| Group | Under 40<br>(% dx\| % cumul) | Under 50<br>(% dx\| % cumul) | Under 60<br>(% dx\| % cumul) | Under 70<br>(% dx\| % cumul) | Under 80<br>(% dx\| % cumul) | Total<br>(% dx\| % cumul) |
| --- | --- | --- | --- | --- | --- | --- |
| PKD1 LoF | 50.6 \| 31.4 | 72.7 \| 45.2 | 90.9 \| 56.5 | 98.7 \| 61.3 | 100 \| 62.1 | 100 \| 62.1 |
| PKD2 LoF | 17.9 \| 11.6 | 46.4 \| 30.2 | 78.6 \| 51.2 | 92.9 \| 60.1 | 100 \| 65.1 | 100 \| 65.1 |
| non-carrier | 8.75 \| 3.49 | 29.2 \| 11.6 | 62.9 \| 25.1 | 88.4 \| 35.2 | 99.5 \| 39.7 | 100 \| 39.8 |

