## Supplemental Figures for "Hypertension increases PPV for polycystic kidney disease in *PKD1* and *PKD2* variant carriers"

***Figure 1.*** (A) Age of enrollees in UKB cohort divided into *PKD1* LoF, *PKD2* LoF and non carriers. (B) Age at hypertension diagnosis. (C) Age at kidney disease diagnosis. (D) Age at end-stage renal disease diagnosis. (E) Empirical cumulative density function (ECDF) of ages at diagnosis for (left) hypertension, (middle) kidney disease and (right) end-stage renal disease.

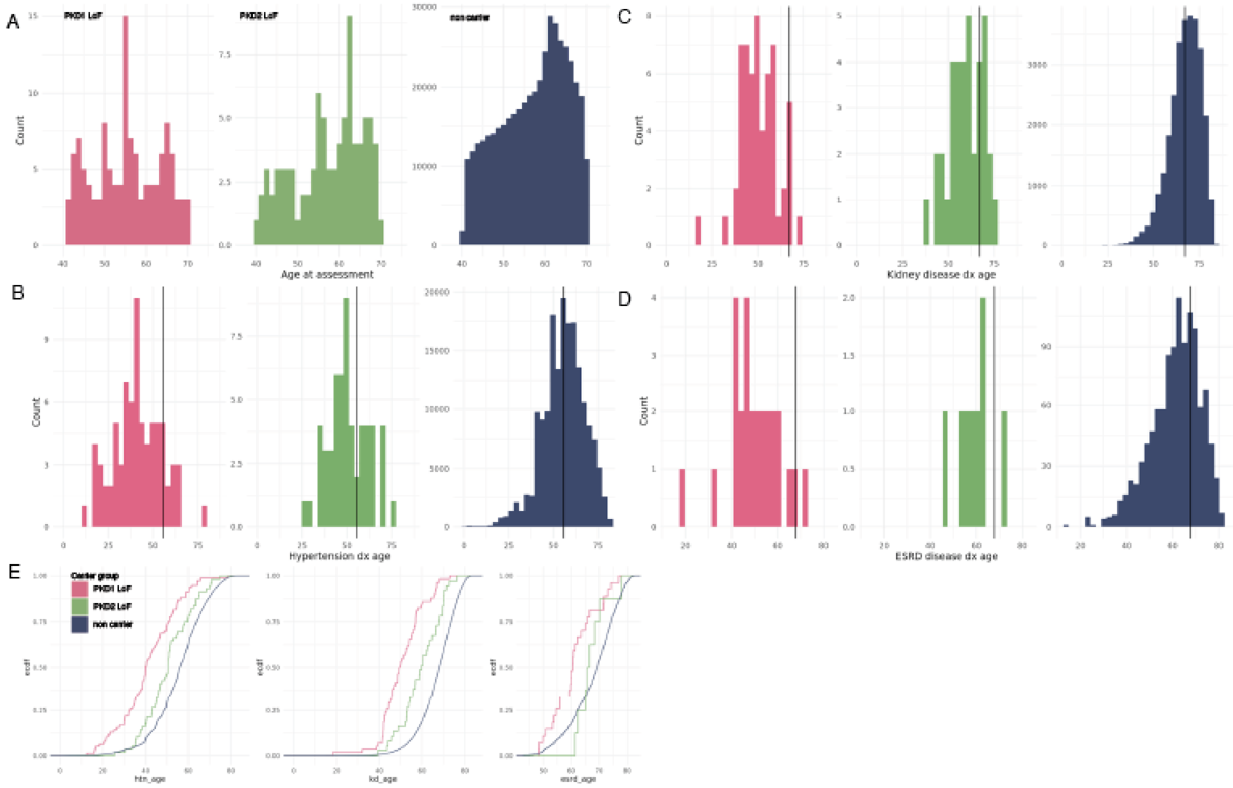

*Figure 2.* Age at incidence of disease for hypertension (right) and kidney disease (left) divided by whether they receive treatment for hypertension or not (UKB fields 6153, 6177).

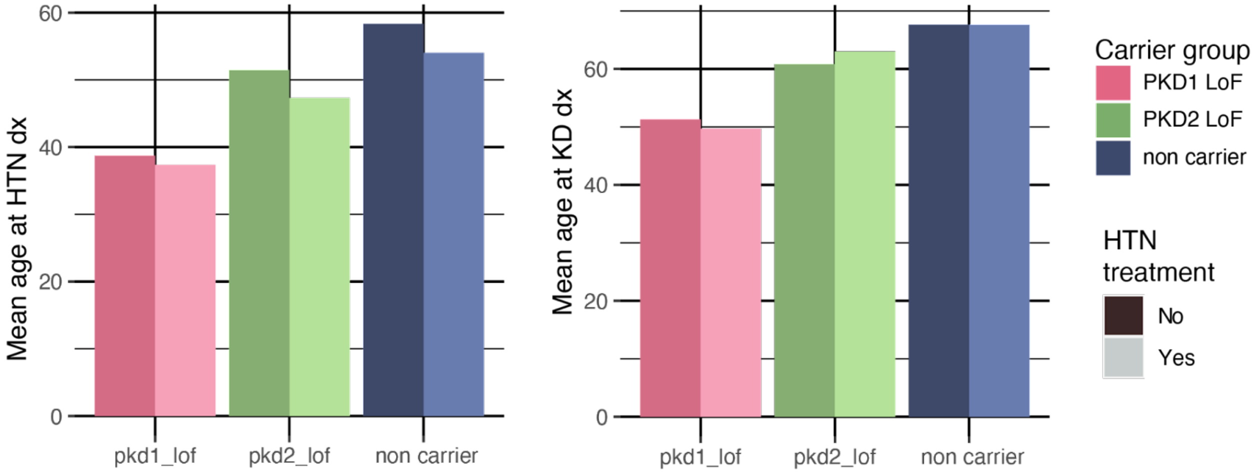


`

*Figure 3.* (A) Time to event (kidney disease diagnosis) for only individuals with a diagnosis of **hypertension**. (B) Time to event (kidney disease diagnosis) for only individuals with a diagnosis of **kidney disease**. (C) Time to event (end-stage renal disease diagnosis) for only individuals with a diagnosis of kidney disease.

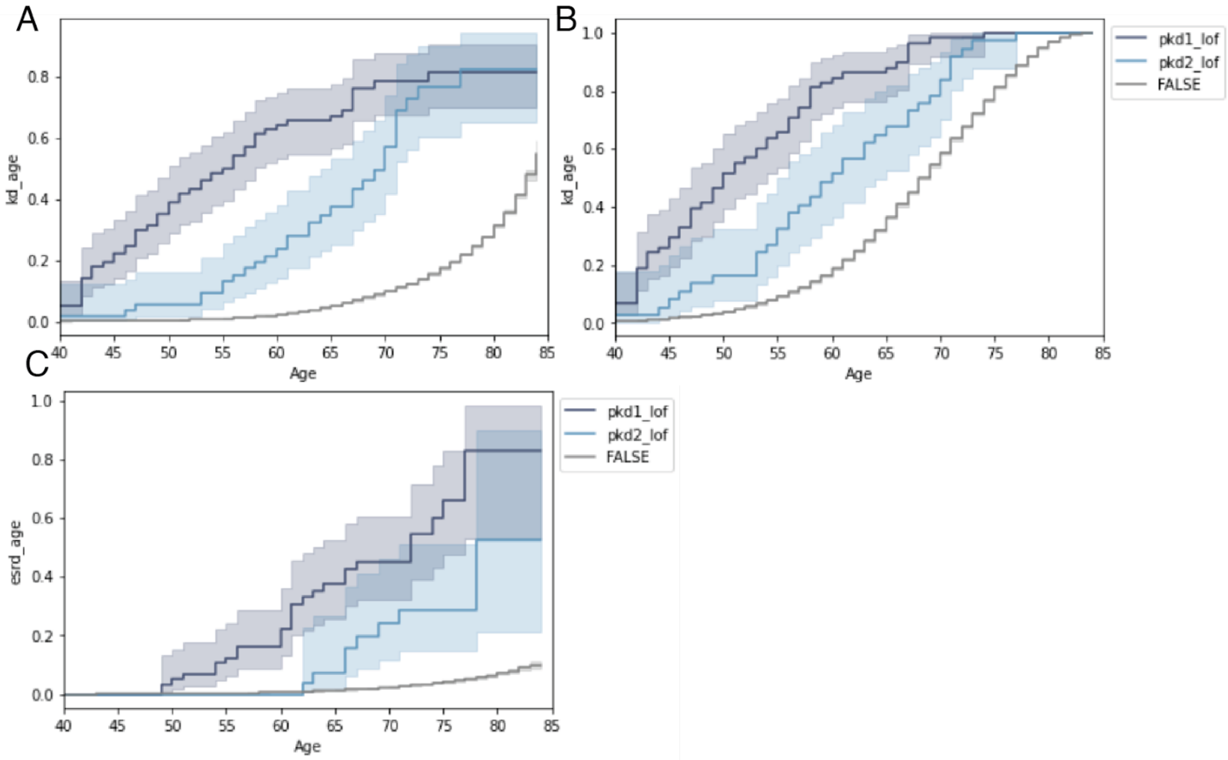

*Figure 4.* eGFR for all individuals, split by prospective/retrospective case status for hypertension as well as kidney disease.

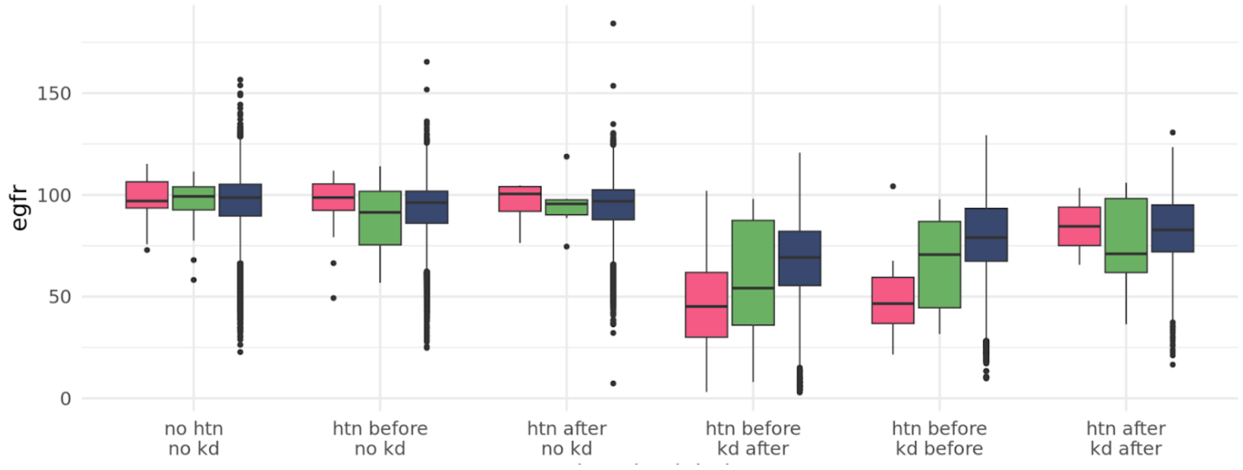

*Figure 5.* Proportion of individuals with either *PKD1* or *PKD2* LoF variants in individuals diagnosed with hypertension by or before a given age.

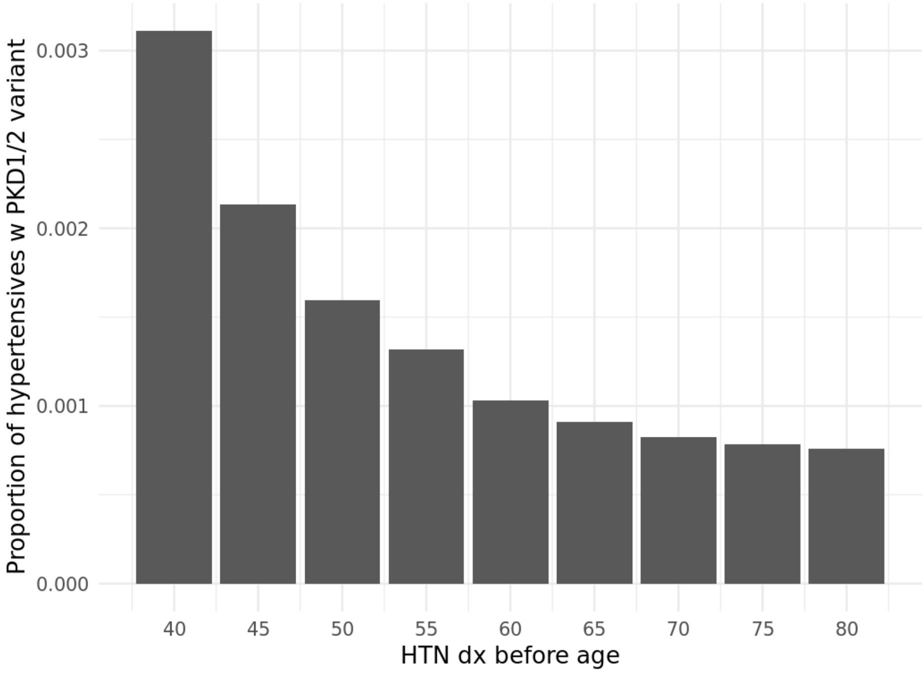
